# PROTOCOL: A cohort study examining the association between mirtazapine and mortality risk in adults with a diagnosis of depression

**DOI:** 10.1101/2021.02.08.21250305

**Authors:** Rebecca M Joseph, Ruth H Jack, Richard Morriss, Roger David Knaggs, Chris Hollis, Julia Hippisley-Cox, Carol Coupland

**Author notes:** **Corresponding author:** Rebecca M Joseph.

## Abstract

This protocol describes a cohort study comparing the risks of mortality and serious self-harm (suicide or near-fatal deliberate self-harm) between adults with depression prescribed mirtazapine, a selective serotonin reuptake inhibitor (SSRI), amitriptyline, or venlafaxine. The study is set within English primary care electronic health records from the Clinical Practice Research Datalink (CPRD) and covers the period 01 January 2005 – 30 November 2018. The study described uses an active comparator new user design: patients are included if they are first prescribed an SSRI before being prescribed mirtazapine, a different SSRI, amitriptyline, or venlafaxine as their second antidepressant. Patients are followed from the initial prescription for the second antidepressant until an outcome (mortality, serious self-harm), end of CPRD follow-up, or study end. Inverse probability of treatment weighting is used to account for confounding variables. The protocol was submitted to the CPRD Independent Scientific Advisory Committee for review and was approved in November 2019 (protocol number 19_241).

## A. Study Title

A cohort study examining the association between mirtazapine and mortality risk in adults with a diagnosis of depression

## B. Lay Summary

Mirtazapine is a treatment for depression in adults. Some studies have found a link between mirtazapine and increased risk of death compared to other antidepressants. However, other factors may make this link seem stronger than it is. For example, doctors may prescribe mirtazapine to terminally ill patients as it can increase appetite and promote sleep.

The aim of this study is to better understand the relationship between mirtazapine and risk of death. We will look at the risk of specific causes of death, including heart disease and cancer. We will also look at whether there is a link between mirtazapine and attempted suicide.

We will use data from the Clinical Practice Research Datalink (CPRD) for this study. The dataset will include anonymised electronic medical records from general practices and hospitals, as well as anonymised death records. We will compare a group of patients taking mirtazapine with groups of patients taking other antidepressants. The groups will be made up of adult patients with depression. We will compare the risk of death or attempted suicide between the groups. Risk factors such as age, sex, and other health conditions will be taken into account in the analyses.

The study will provide further information about whether there is a link between mirtazapine and risk of death or attempted suicide. This will include whether there is a link with particular causes of death. This will be important information for doctors and patients when considering suitable antidepressant treatment.

## C. Technical Summary

Previous studies have found an association between the antidepressant mirtazapine and increased risk of mortality. Further research is needed to confirm this association and to examine specific causes of death. The proposed study will compare mirtazapine to other antidepressants and investigate cause-specific mortality, controlling for relevant confounding.

We will perform a cohort study using the active-comparator new user design, selecting patients prescribed antidepressants between 1st January 2005 and 30th November 2018. Linked CPRD primary care data, Office for National Statistics (ONS) mortality data, and Hospital Episode Statistics (HES) admitted patient care data will be used.

The study population will comprise adult patients diagnosed with depression whose first antidepressant was a selective serotonin reuptake inhibitor (SSRI) and who were subsequently prescribed mirtazapine, venlafaxine, amitriptyline, or a different SSRI. Follow-up will start from the date of starting the new antidepressant.

There will be five exposure groups: those who switched to mirtazapine, those who started mirtazapine while continuing the original SSRI, and those who switched to venlafaxine, amitriptyline, or a different SSRI.

The outcomes will be all-cause and cause-specific mortality, and suicide/near-fatal deliberate self-harm. Cause-specific mortality will include cardiovascular disease, cancer, and the most common causes of death found within the study population. Date and cause of death will be taken from the ONS mortality dataset while near-fatal deliberate self-harm will be defined using a combination of primary care and HES data.

We will account for confounding factors including demographic characteristics, lifestyle factors, drug exposures, and comorbidities. These will be used to estimate propensity scores for each of the comparator drugs compared with mirtazapine.

Baseline characteristics and the most frequent causes of death will be summarised. Survival analysis using Cox and Fine-Gray (competing risk) regression will be performed, accounting for propensity scores.

## D. Outcomes to be Measured

All-cause mortality; cause-specific mortality (including cardiovascular disease and cancer); suicide or near-fatal deliberate self-harm.

## E. Objectives, Specific Aims and Rationale

Objective: to examine the association between the antidepressant mirtazapine and risks of mortality and suicidal behaviour by exploring specific causes of death in a cohort of patients with depression. Mortality rates for patients taking mirtazapine and patients taking other antidepressants will be compared.

### Specific aims

1. To describe and compare the characteristics of patients prescribed mirtazapine compared to alternative antidepressants
2. To estimate rates of all-cause mortality, cause-specific mortality, and suicide/near-fatal deliberate self-harm in those prescribed mirtazapine and alternative antidepressants
3. To test the hypothesis that the rate of all-cause mortality differs between those taking mirtazapine and those taking alternative antidepressants
4. To test the hypotheses that rates of specific causes of death differ between those taking mirtazapine and those taking alternative antidepressants using survival analysis with competing risks
5. To test the hypothesis that the rate of suicide/near-fatal deliberate self-harm differs between those taking mirtazapine and those taking alternative antidepressants using survival analysis with competing risks

### Rationale

mirtazapine has been associated with an increased risk of all-cause mortality(1). This association could reflect a direct effect of the drug (e.g. increased risk of suicide) or confounding factors that influence choice of antidepressant. This study will generate additional evidence about the comparative risk of mortality, cause-specific mortality, and suicide/near-fatal deliberate self-harm in patients taking mirtazapine compared with other antidepressants. Differences between the treatment groups will be summarised and accounted for in analyses.

## F. Study Background

This study will examine the association between mirtazapine, mortality, and self-harm in patients with depression. Mirtazapine is licensed to treat depression in adults and has similar efficacy to other antidepressants(2). The tolerability of mirtazapine is also similar to other antidepressants, although the typical side effects differ(2,3). Mirtazapine was the 7^th^most frequently prescribed antidepressant between 2000-2011 in a UK cohort of patients with depression(4). Openly-available English prescribing data(5) show the number of mirtazapine prescriptions is increasing over time, with 8,938,362 prescriptions issued in the community in 2018 compared with 2,437,500 in 2008(6).

Observational research set within UK primary care has highlighted an increased risk of all-cause mortality in patients prescribed mirtazapine compared with other antidepressants(1,7). These studies also demonstrated an increased risk of suicide and attempted suicide/self-harm associated with mirtazapine(1,4). While the increased mortality could reflect this increased risk of suicide, additional factors may also contribute. As different antidepressants have similar efficacy, side effects and individual patient characteristics may influence choice of antidepressant(3). If so, factors influencing choice of antidepressant may confound the relationship between mirtazapine and mortality.

Clinicians may preferentially prescribe mirtazapine to certain patient populations because of its positive effects on appetite(8–10) and sleep(11). For instance, the British National Formulary(12) suggests prescribing mirtazapine to patients with sleep disturbance in depression (chapter 4.1.1). It is also possible mirtazapine is preferentially prescribed to treat depression in patients on the palliative care pathway(13–16). Studies need to account for the possibility of such confounding.

To better understand the association between mirtazapine and mortality it is also necessary to explore specific causes of death. As described above, mirtazapine may be associated with increased risk of suicide or self-harm. In addition, as mirtazapine has been linked to weight gain there could be an association between mirtazapine and deaths due to cardiovascular disease. This study will explore the risk of deaths due to suicide, cardiovascular disease, cancer, and other common causes of death within the study population.

UK guidelines for the treatment of depression(17) recommend that, where antidepressants are prescribed, SSRIs should be prescribed in the first instance. If ineffective or not tolerated, the guidelines recommend switching to a “different SSRI or a better tolerated newer-generation antidepressant”. If the patient is willing to tolerate the increased side-effect burden, augmenting treatment with other drugs including mirtazapine could be considered. However, where patients have another chronic physical health problem mirtazapine may be preferred as the first antidepressant over SSRIs(18). Therefore, patients whose first antidepressant is mirtazapine are likely to differ systematically from patients whose first antidepressants are SSRIs. For this reason, the current study will include patients who switch from SSRIs to mirtazapine or one of the comparator drugs (see below) rather than new users.

Those who switch to mirtazapine will be compared to those who switch to a different SSRI, or to venlafaxine or amitriptyline. The SSRI group are included as the standard treatment group, if prescribing according to guidelines. We are planning to include SSRIs as a single group to avoid reducing the sample size for the other comparison groups. There is evidence that different SSRIs have similar risk/benefit profiles, including similar risk of mortality(1). The venlafaxine group are included to represent an alternative “newer-generation antidepressant”. Amitriptyline, a tricyclic antidepressant, is included as a comparison group as previous studies have found an increased risk of mortality similar to mirtazapine(1). A final comparison will be made between those who switch to mirtazapine and those who augment SSRI treatment with mirtazapine. The study will be therefore able to compare many of the common treatment options available for treatment of depression.

The association between mirtazapine and mortality was highlighted in studies using the QResearch database(1,4,7). A number of CPRD studies have previously investigated antidepressant safety. For example, studies report a decreased risk of myocardial infarction associated with SSRIs(19) and decreased risk of certain cancers associated with tricyclic antidepressants(20). However, many studies grouped antidepressants by antidepressant class, with few reporting results for individual drugs. Studies reporting results for mirtazapine demonstrated an increased risk of significant weight gain(10), and a possible increased risk of seizures(21).

## G. Study Type

Descriptive (aims 1 and 2) and hypothesis testing (aims 3 - 5).

## H. Study Design

Cohort study using the active-comparator new user design(22). Adult patients initially taking an SSRI for depression who switch to mirtazapine or one of the comparator drugs (see Section N) will be followed from the date of switch until the end of drug exposure, starting a different antidepressant, death, leaving the cohort, or end of study period. The outcomes will be all-cause mortality, cause-specific mortality, and suicide/near-fatal deliberate self-harm. We will generate propensity scores using potential confounding variables and account for these in analyses. We will calculate crude and age-sex adjusted mortality rates and use survival analysis to estimate the risk of mortality associated with the different antidepressants compared to mirtazapine.

### This design will

- Reduce the scale of unmeasured confounding by ensuring exposure groups are more similar at index date
- Avoid potential immortal time bias by defining an equivalent index date across exposure groups
- Allow the relative timing of events to be taken into account, defining confounders using information recorded prior to exposure to the drug of interest
- Compare mirtazapine exposure to alternative antidepressants rather than a ‘no use’ category, providing results that are more clinically meaningful

Mirtazapine is not recommended as the first drug treatment for depression in adult patients: guidelines recommend initially trying an SSRI. It is possible that patients whose first antidepressant is mirtazapine are systematically different from patients initiating an SSRI as recommended. For this reason we have chosen to start follow-up at the first switch from SSRI, rather than first ever antidepressant. We explored the numbers of patients initiating mirtazapine or the comparator drugs and found the counts were similar to the numbers who switched.

We are planning to repeat the analysis in QResearch, given appropriate approvals and agreements. If successful we will combine the results by meta-analysis as in previous studies (e.g. Vinogradova et al. 2019 (23)).

## I. Feasibility Counts

We extracted the prescription dates for SSRIs, mirtazapine, amitriptyline, and venlafaxine from the September 2019 CPRD dataset release. After applying the acceptability and follow-up date criteria described in section L, we generated the following sample size estimates:

– First antidepressant is an SSRI and at least one subsequent prescription for
– mirtazapine (n=40,200)
– venlafaxine (n=9492)
– amitriptyline (n=55,301)
– a different SSRI (n=135,702)

The dataset will be limited to those with linkage to the ONS Mortality and the HES datasets. Assuming approximately 50% are linked, new estimates are:

- switch to mirtazapine: n=20,000
- switch to venlafaxine: n=4700
- switch to amitriptyline: n=27,500
- switch to a different SSRI: n=67,800

These represent crude estimates and do not account for factors such as evidence of depression, the length of time between prescriptions, or any exposure to other antidepressants.

## J. Sample Size Considerations

We estimated the power of detecting a difference in all-cause mortality and in the rate of suicide/near-fatal deliberate self-harm between patients switching to mirtazapine compared to patients switching to each of the comparator drugs. Estimated rates were based on rates published by Coupland et al (all-cause mortality(1), suicide/attempted suicide/self-harm(4)), based on cohorts of patients aged 20-64 years old with a diagnosis of depression.

The following Stata command was used to calculate power, based on the estimated sample sizes from Section I:

“power twoproportions r1 r2, alpha(0.05) n1(N1) n2(N2)”

where r1 is the incidence rate for the comparator drug, r2 is the incidence rate for mirtazapine, N1 is the sample size for the comparator drug, and N2 is the sample size for mirtazapine. This calculation estimates power to detect a difference between two proportions therefore does not take length of follow-up into account.

### Crude mortality rates

- Mirtazapine 110 deaths / 10,343 person years = 0.0106 (106 per 10,000 person years)
- Venlafaxine 73 deaths / 15,835 person years = 0.0046 (46 per 10,000 person years)
- Amitriptyline 194 deaths / 19,845 person years = 0.0098 (98 per 10,000 person years)
- SSRIs 990 deaths / 228,233 person years = 0.0043 (43 per 10,000 person years)

### The crude rates of suicide/attempted suicide/self-harm

- Mirtazapine 16 events / 10,343 person years = 0.0015 (15 per 10,000 person years)
- Venlafaxine 13 events / 15,835 person years = 0.0008 (8 per 10,000 person years)
- Amitriptyline 4 events / 19,845 person years = 0.0002 (2 per 10,000 person years)
- SSRIs 79 events / 228,233 person years = 0.0003 (3 per 10,000 person years)

### Estimated power, all-cause mortality

- venlafaxine vs mirtazapine: power = 0.992
- amitriptyline vs mirtazapine: power = 0.140
- SSRIs vs mirtazapine: power = 1.00

### Estimated power, suicide/self-harm

- venlafaxine vs mirtazapine: power = 0.169
- amitriptyline vs mirtazapine: power = 0.997
- SSRIs vs mirtazapine: power = 0.998

Based on the estimated counts and rates, the study is likely to have sufficient power for many of the comparisons. Comparisons with a smaller risk difference may be underpowered (e.g. mirtazapine vs amitriptyline for mortality, mirtazapine vs venlafaxine for suicide/near-fatal deliberate self-harm). As discussed in Section I, the feasibility counts are likely to be overestimates. Power will be discussed in the planned publication, and confidence intervals will be presented with all estimates.

If the final number of patients in the venlafaxine group is low, we will consider using a combine selective-noradrenaline reuptake inhibitor (SNRI) group including venlafaxine and duloxetine.

## K. Planned use of Linked Data

Date and cause of death will be determined using the ONS mortality dataset. The definition of near-fatal deliberate self-harm will include data from the HES admitted patient dataset. The analysis will account for socio-economic status using the Townsend Score from the deprivation dataset. The cohort will be restricted to patients at practices contributing to the ONS mortality dataset and HES admitted patient care dataset. Follow-up will be censored at the end of the linkage window.

## L. Definition of the Study Population

Preliminary code lists to be used throughout study definition are available from the authors, with further details in Appendix 1. A diagram explaining the time windows described below has also been provided as an appendix (Appendix 2).

Only data recorded after the up-to-standard date (an indicator of data quality provided by CPRD) will be included in the study. All patients will require at least 1 year of follow-up after this up-to-standard date or joining the practice.

### Study window

1^st^January 2005 – 30^th^November 2018; this period begins the year after the patent for mirtazapine expired (2004), when generic versions became available, and ends at the end of the current HES linkage window

### Eligibility window start

latest of up-to-standard date + 1 year, current registration date + 1 year, first registration date + 1 year, date of turning 18 years old, 1^st^January 2005

### Eligibility window end

earliest of transfer out date, last collection date, death date, prescription for a third antidepressant, 30^th^November 2018

### First SSRI

date of first prescription for an SSRI during the eligibility window

### Index prescription

first prescription for mirtazapine or one of the comparator antidepressants (see Section N) recorded after the first SSRI; index date is the date of the index prescription

### Inclusion criteria

- Permanently registered acceptable patient <u>AND</u>
- Registered at a practice linked to the ONS mortality dataset and HES admitted patient datasets <u>AND</u>
- At least 0 days of follow-up (eligibility start <= eligibility end) <u>AND</u>
- First prescription for an SSRI recorded after the start of the eligibility window <u>AND</u>
- At least 1 prescription for mirtazapine or the comparator drugs within the eligibility window <u>AND</u>
- Index prescription is recorded after the first SSRI and either during exposure to or less than 90 days after the end of exposure to the first SSRI <u>AND</u>
- A depression Read code is recorded less than 12 months before the first SSRI prescription up to the index prescription

### Exclusion criteria

- Age 100 years old or above at index date <u>OR</u>
- A prescription for any antidepressant other than an SSRI prior to the index prescription <u>OR</u>
- Any previous antidepressant switch prior to index prescription <u>OR</u>
- A record of bipolar disorder or schizophrenia prior to index prescription

### Follow-up

from date of index prescription to death, starting another different antidepressant, end of index antidepressant use (see Section N), or end of eligibility

## M. Selection of Comparison Groups

Patients will be grouped according to the antidepressant prescribed at the index prescription. The exposure and comparison groups will be defined as described in Sections L and N. Each comparison group will have an equivalent start point, i.e. patients have to survive until starting their index antidepressant.

## N. Exposures, Outcomes and Covariates

### Exposure

Exposure is defined as a prescription for mirtazapine or one of the comparator antidepressants. The antidepressant prescribed at the index prescription will determine which exposure groups patients are in. The comparator antidepressants will be: venlafaxine, amitriptyline, and a different SSRI. A further comparison will be made between those who switch to mirtazapine and those who augment SSRI treatment with mirtazapine (i.e. those who continue to be prescribed the original SSRI after starting mirtazapine).

As described in Section J, if numbers of patients switching to venlafaxine are low we will consider using a combined SNRI group (venlafaxine and duloxetine). If numbers allow, we will group those in the augmented treatment cohort according to the initial SSRI and perform a subgroup analysis to investigate possible interactions between SSRIs and mirtazapine.

The period of exposure to a drug will be calculated based on prescription records. The start (typically the prescription date) and stop date of each individual prescription will be calculated using a published drug data preparation algorithm(24). We will consider the exposure period as continuous where prescription start and stop dates overlap.

To define the ‘period-at-risk’, a six month risk carry-over period will be added to the stop date of individual prescriptions. Periods-at-risk will run from the first prescription for the drug of interest until the earliest of: stopping the drug + the carry-over period, a prescription for a different antidepressant, death, or the end of the eligibility window. The analyses will include only one period-at-risk per patient.

The dose of each prescription will be estimated by dividing the strength by numeric daily dose. Fluoxetine-equivalent doses will be calculated(25).

### Outcomes

All-cause mortality, cause specific-mortality, and suicide/near-fatal deliberate self-harm(26). Cause of death will be based on the underlying cause of death field within the ONS dataset. Date of death will be taken from the ONS dataset. Specific causes of interest are: cardiovascular disease, cancer, and suicide. Other frequent causes of death identified in the dataset will also be analysed. A composite outcome of suicide/near-fatal deliberate self-harm will be defined using the primary care, HES, and ONS mortality data: the first record in any dataset with a relevant code will be used as the event date.

### Covariates

Covariates will be included in the propensity score models. These include likely confounding factors and factors associated with mortality. The latter factors are those included in the Charlson Comorbidity Index(27) and QMortality risk prediction algorithm(28). Covariates are listed below.

#### Demographic characteristics

age in index year (taking 01 Jan to be each patient’s birthday); sex (male/female); ethnicity (most recent code prior to index date); socio-economic status (Townsend score quintile); practice region.

#### Lifestyle indicators

body mass index; smoking status; alcohol use (non-/ former/ occasional/ moderate/ heavy drinker), ‘alcohol misuse’ will also be defined.

#### Medication use

opioid use; glucocorticoid use; non-steroidal anti-inflammatory drugs (NSAIDs); statins; antipsychotics; anxiolytic medication; hypnotic agents.

#### Comorbidities

depression severity (mild/ moderate/ severe); insomnia; loss of appetite; unexplained/ unintentional weight loss; epilepsy/ seizures; anxiety (including panic disorder); personality disorder; eating disorders; Huntington’s disease; Parkinson’s disease; multiple sclerosis; hypertension; myocardial infarction; congestive heart failure; peripheral vascular disease; cerebrovascular disease; angina; atrial fibrillation; venous thromboembolism; hemiplegia; dementia; chronic obstructive pulmonary disease (COPD); asthma; dyspnoea; rheumatologic disease; peptic ulcer disease; diabetes mellitus (type 1 and 2), with and without complications; renal disease; cancer; metastatic solid tumour; liver disease, mild and moderate; acquired immune deficiency syndrome (AIDS); leg ulcer; pancreatitis; poor mobility; anaemia; intellectual disability; unplanned hospital admission; and living in a care home.

Presence of covariates will be assessed using data recorded on or before the index date. To define depression severity, the depression Read codes have been categorised according to severity, where possible. Patients will be assigned the highest severity in their record prior to index date. Missing values will be imputed. To be classified as a medication user the patient will require a prescription for that drug within the 6 months prior to index date. For ethnicity, body mass index, smoking status, and alcohol use the most recent record prior to index date will be used. Medical conditions will be considered present at baseline if there is a record of that condition prior to index date. For cancer, ‘recent’ (record in past year) and ‘ever’ categories will be used.

### Other variables

A series of sensitivity analyses excluding patients with prior records for palliative/end-of-life care, near-fatal intentional self-harm, bipolar disorder, and schizophrenia will be performed.

### Code lists

Where possible, code lists have been sourced through publications, clinicalcodes.org, or the CALIBER phenotype resource(29). In some cases, new preliminary code lists have been developed which will be refined by clinicians in the research team.

Antidepressants including mirtazapine: individual products were identified by searching by generic and brand names in the product dictionary; categories are then based on the British National Formulary (BNF)(12) chapters.

Other medication code lists were based on the recorded BNF chapters: antipsychotics (chapters 4.2.1 and 4.2.2), anxiolytic medications (chapter 4.1.2), hypnotic agents (chapter 4.1.1), NSAIDs (chapter 10.1.1), statins (chapter 2.12.4), and opioids (chapter 4.7.2).

Causes of death codes are based on ONS short list of cause of death(30). ICD-10 codes for pre-specified causes of interest are C00-C97 (malignant neoplasms); I00-I99 (diseases of the circulatory system); X60-X84, Y10-Y34 (intentional self-harm/ event of undetermined intent). The ONS short list will be used to categorise additional causes of death in the dataset.

For depression, a new list created by combining Read code lists from three sources: the Quality and Outcomes Framework depression cluster (QOF v38), a list published by John et al(31), and a composite list published by Carreira et al(32). In creating the list we favoured specificity over sensitivity. First we selected all codes included in the QOF cluster plus any equivalent codes in the other list. From these, we retained only the codes most specific to major depression, dropping codes with additional mental health conditions (e.g. “bipolar affective disorder, now depressed”).

Palliative/end-of-life care: a new Read code list was developed by the study team.

Self-harm: the current Read code list is based on codes used by Carr et al (33) and Coupland et al(4). ICD-10 codes for intentional self-harm/event of undetermined intent (X60-X84, Y10-Y34) will be used to find records in the HES data. These code lists will be refined to meet the definition of near-fatal deliberate self-harm described in Douglas et al(26).

## O. Data/ Statistical Analysis

1. Summary of baseline characteristics of patients according to exposure group. Continuous variables will be summarised using the median and inter-quartile range. Categorical variables will be summarised using counts and percentages. The comparison groups will be compared to the mirtazapine-switch group using appropriate difference tests (chi-squared or Mann-Whitney U tests).
2. Length of follow-up and reasons for end of follow-up (e.g. outcome, stopping the drug of interest, starting a new antidepressant) for each of the exposure groups will be summarised.
3. Tabulation of the most frequent causes of death according to exposure group.
4. Calculation of mortality/incidence rates for the different exposure groups. These rates will be age-sex standardised to the whole study population. 95% confidence intervals will be reported. Mortality/incidence rates will be calculated for all causes of death, the pre-specified causes of death, the most common causes of death identified in step 2, and suicide/near-fatal deliberate self-harm.
5. Missing values will be imputed (Section Q) and propensity scores estimated for each of the imputed datasets. The analyses will be run on each of the imputed datasets, accounting for each propensity score, before the results are combined (the ‘within’ approach described by Granger et al.(34)). Propensity scores will be generated using a multinomial regression model including the listed covariates (potential confounding variables and risk factors for the outcome). Appropriate model diagnostics will be performed to ensure correct specification of the propensity score and to evaluate the overlap of the distribution between exposed and groups(35). The inverse probability of treatment weighting will be used to account for propensity scores in the outcome models(36).
6. The risk of all-cause mortality associated with exposure to mirtazapine vs. the comparator antidepressants will be estimated using Cox regression. The outcome will be time to death, censoring when patients leave the cohort. A simple model adjusting for age and sex will be performed, followed by a model adjusting for propensity scores. The proportional-hazardsN assumption will be tested for all models and appropriate adjustments made where necessary.
7. Cause-specific mortality will be studied using Fine-Gray regression to account for competing risks. The steps described in step 5 will be repeated for this analysis.
8. Fine-Gray regression will be used to study the risk of suicide/near-fatal deliberate self-harm, including death from another cause as a competing risk. The steps described in step 5 will be repeated for this analysis.
9. The effect of antidepressant dose will be explored for starting dose and median dose, for example by including dose as a covariate or stratifying according to high/low starting dose.
10. As mentioned in Section H, we plan to repeat the study using the QResearch database, given appropriate approvals. The data preparation and analysis will be repeated as closely as possible and, if appropriate, results will be combined using meta-analysis techniques as in previous studies (e.g. Vinogradova et al. 2019 (23)).

### Sensitivity analyses

Sensitivity analyses excluding patients with a prior record for palliative/end-of-life care, near-fatal deliberate self-harm, bipolar disorder, or schizophrenia will be performed.

All-cause mortality rates and age-sex-adjusted Cox regression models will be repeated after a) excluding patients who have a record of palliative/end-of-life care prior to index date, and b) excluding patients with a record of self-harm prior to index date.

Subgroup analyses will be performed including those aged 18-64 years and those aged 65-99 years at index date.

The analyses will be repeated using a shorter risk carry-over period (30 days after prescription stop date). The longer carry-over period in the main study (6 months) is included to allow for potential longer-term effects of antidepressants, such as weight gain, but may introduce misclassification bias. The 30-day carry-over period will reflect the immediate effects of the drug and shorter-term withdrawal effects.

Analyses will also be repeated after restricting the maximum follow-up length to one year after index date. It is possible the antidepressants may have different tolerability and effectiveness in the study cohort, leading to systematic differences in how long patients persist on the different drugs.

If numbers allow, we will investigate possible drug interactions within the augmented treatment group by further grouping patients according to the initial SSRI and comparing outcome rates.

### Multiple testing

A significance level of 0.05 will be used throughout the analyses. Where possible, 95% confidence intervals will be reported alongside estimates. The significance level will be made clear in publications and the results interpreted with appropriate caution.

### Disclosure control

Disclosure control techniques recommended by the ONS will be used to minimise the risk of reidentification. Minimum cell counts of 5 will be used.

## P. Plan for Addressing Confounding

Propensity score adjustment will be used to address confounding, as described in Section O. The study population will also be restricted to patients with a depression Read code with similar antidepressant treatment histories, reducing the risk of indication biases. Although imperfect, this step will reduce the level of unmeasured variation by making the cohort members more similar at baseline(37).

## Q. Plans for Addressing Missing Data

Multiple imputation by chained equations will be used to impute missing values for baseline covariates (body mass index, ethnicity, socio-economic status, alcohol use, smoking status, depression severity). Twenty datasets will be imputed and the results will be combined using Rubin’s rules. All variables included in the propensity score models and outcome models, including survival time, death, and cause of death, will be included in the imputation models. Drug exposure and medical condition variables will be assumed not present if not recorded. The amount of missing data will be reported in the study outputs.

## R. Patient and Public Involvement

The study team includes two patient and public involvement (PPI) representatives from the MindTech Involvement team (https://www.mindtech.org.uk/involvement), a group of individuals with lived experience of mental health conditions. The PPI representatives have helped develop the protocol and will attend project meetings and additional meetings throughout the analysis. The project will therefore have patient representation in study design, analysis, interpretation, and dissemination of results (including co-authorship of papers). We will also discuss the progress and results of the study at meetings of the Involvement Team.

## S. Plans for Disseminating Study Results

We will publish the study in a peer-reviewed journal article and follow the reporting guidelines of the RECORD statement(38,39) (an extension of the STROBE guidelines). We will submit abstracts for presentation at conferences.

## T. Possible Limitations

There is a risk of unmeasured confounding, confounding by indication, and channelling bias in this study. We attempt to address this in part by restricting the study population to those with a Read code for depression, starting follow-up at the first antidepressant switch (or addition, for mirtazapine), and calculating propensity scores to account for systematic differences between exposure groups at baseline. This will reduce some of the heterogeneity within the population and ensure groups are more similar. However, the possibility of unmeasured confounding will still be present. We will acknowledge this when interpreting the results of the study.

Restricting the study population will reduce the generalisability of the findings. For instance, we are restricting the cohort to patients with a diagnosis of depression first prescribed antidepressants after the age of 18 years old. As the risk of confounding is high, we have focused on reducing the potential impact of this at the cost of generalisability. As major depression is the only licensed indication for mirtazapine, and mirtazapine is not licensed for use in under 18 year-olds, we may not in fact exclude many patients exposed to our main exposure of interest. We will quantify the numbers of patients dropped at each stage of cohort creation and discuss generalisability in our study outputs.

There is likely to be measurement error/ misclassification in our exposure variable. We are using prescriptions as a proxy for drug use. Non-adherence is a recognised problem for many prescription medicines. Furthermore it is possible antidepressants are prescribed in secondary care, thus we may underestimate true exposure. The likelihood of this is difficult to quantify, although in adult patients it is probable the majority of antidepressants are initiated in primary care, or continued in primary care if initiated in secondary care. These errors may introduce misclassification bias, the direction of which is difficult to predict. This problem affects all pharmacoepidemiology studies using routine data and will be acknowledged and discussed in study outputs.

The study may lack power if the number of events is low, for instance for some specific causes of death. We will present confidence intervals for estimates and discuss the possible impact during interpretation of results.

## Data Availability

The preliminary code lists submitted with the protocol are available on request from the authors. Final code lists will be shared as part of future publications.

## Funding and acknowledgments

We thank our patient representatives, Debbie Butler and Dave Waldram, for their advice in the development of this protocol.

This work has been funded by the National Institute for Health Research (NIHR). The authors are supported by the NIHR Nottingham Biomedical Research Centre. RM is supported by the NIHR MindTech MedTech and In Vitro Fertilisation Collaboration and the NIHR Applied Research Collaboration East Midlands. The views represented are the views of the authors alone and do not necessarily represent the views of the Department of Health in England, NHS, or the National Institute for Health Research.

## Appendix 1 Code Lists

The preliminary code lists submitted with the protocol are available on request from the authors. The final code lists will be shared as part of future publications. Key resources used to develop code lists include the CALIBER phenotyping resource (https://www.caliberresearch.org/portal/phenotypes/chronological-map) and the ClinicalCodes repository (https://clinicalcodes.rss.mhs.man.ac.uk/).

## Appendix 2 Time Windows

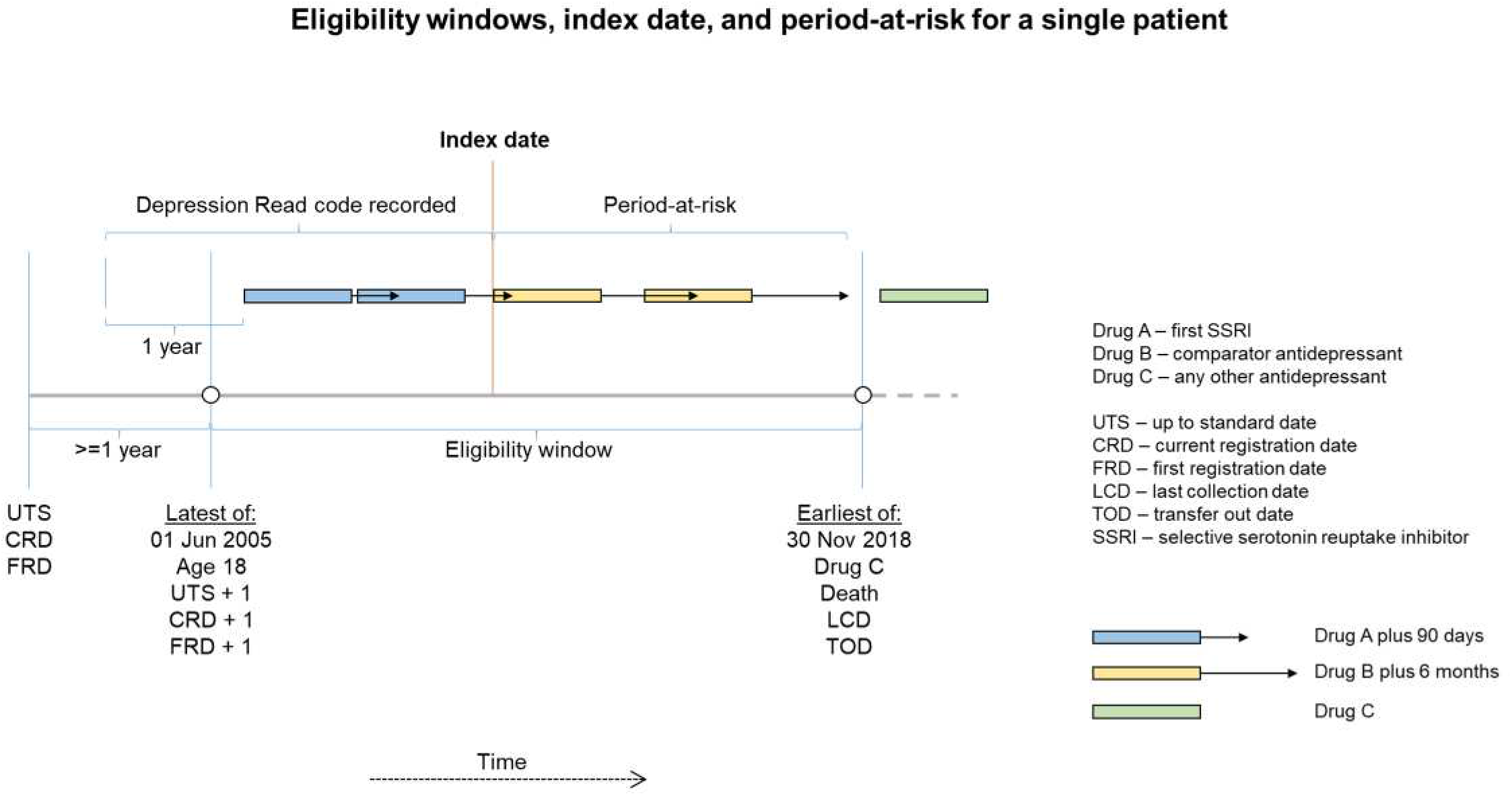

